# Cost Reduction and Care Quality in Resource-Limited Settings – A Protocol for An Umbrella Review of Sterile versus Examination Gloves for Wound Care

**DOI:** 10.1101/2025.05.21.25328093

**Authors:** Cyril E. Dzah, Joy G. L. Bulley, Valentine K. Dzah

**Affiliations:** Adidome Government Hospital, Adidome, Ghana (Ghana Health Service); Volta Regional Hospital, Hohoe, Ghana (Ghana Health Service)

**Keywords:** Cost Reduction, Care Quality, Resource-Limited Setting, Umbrella Review, Glove Choice

## Abstract

**Introduction:** Sterile gloves are widely used in wound care due to their perceived protection against infection. However, the high cost of sterile gloves poses challenges in resource-limited settings. On the other hand, examination or clean gloves (non-sterile) are more affordable and easily accessible but are often perceived as less protective against infection. This umbrella review aims to compare the use of sterile versus examination gloves for wound care in terms of care quality (protection against infection) and cost, and explore the implications for healthcare delivery in low-resource environments.

**Methods and Analysis:** This study will be conducted as an overview of systematic reviews. A comprehensive search of PubMed, CINAHL and MEDLINE will be undertaken to identify relevant systematic reviews with meta-analyses comparing sterile and examination gloves in wound care and or minor surgical procedures. Data will be extracted and synthesized narratively. The AMSTAR 2 tool will be used for quality appraisal of included systematic reviews.

**Ethics and Dissemination:** As this study involves secondary data from published systematic reviews, ethical approval will not be required. The findings will be published in a peer-reviewed journal and presented at relevant global health and clinical care conferences.

**PROSPERO Registration Number:** **CRD420251051136**

**Strengths and Limitations of This Study:**

- This study will provide a high-level synthesis of existing systematic reviews on glove use in wound care and or minor surgical procedures.
- The study will employ a narrative synthesis with subgroup analysis where applicable, to enable a better understanding of findings.
- The study will use the AMSTAR 2 tool to assess the quality of included systematic reviews.
- Results may be limited by variability in outcome reporting and the number of high-quality systematic reviews available.
- The overview will not include primary studies not covered in existing systematic reviews.

## INTRODUCTION

Access to safe, effective, and affordable wound care is crucial for early healing and the overall well-being of patients [1–3]. From treating minor cuts to managing post-surgical wounds, proper dressing techniques help prevent infections and promote wound healing [4,5]. One important aspect of wound care is the type of gloves used during the procedure [6]. Sterile gloves are often preferred as the standard for wound care and minor surgical procedures because of their ability to protect against wound infections [7,8]. However, the use of sterile gloves in many low- and middle-income countries (LMICs) pose significant financial burden to both the health facility and the patient receiving care due to the high cost of the glove [9,10]. As a result, in the quest to meet sterile gloves demand, other aspects of care delivery may be compromised.

To meet the health needs of people without compromising the quality of care, the World Health Organization (WHO) has emphasized the importance of adapting medical guidelines to local realities, particularly in settings where high costs may limit access to quality care [11–13]. In such situations, the goal is to maintain or improve on the level of safety and quality while using more affordable alternatives. Therefore, if examination gloves can provide adequate protection during wound care at a fraction of the cost of sterile gloves, they could offer a valuable solution for under-resourced health systems striving to deliver quality care to all.

Although several systematic reviews have explored the effectiveness of sterile versus examination gloves in wound care and minor surgical procedures, there are limited studies that brought this evidence together in one comprehensive review. This study therefore aims to address that gap by conducting an overview of systematic reviews, bringing together findings from multiple high-quality systematic reviews to provide a clearer and broader understanding. It will evaluate both the care quality (infection prevention) and cost implications of using sterile versus examination gloves, to inform practical and evidence-based decisions in resource-limited settings. The findings will help guide healthcare providers and policymakers in choosing strategies that are both cost-efficient and safe for patients.

## METHODS AND ANALYSIS

### Protocol Development

The protocol for this study follows the Preferred Reporting Items for Systematic Review and Meta-Analysis Protocols (PRISMA-P) guidelines. The protocol is officially registered with the International Prospective Register of Systematic Reviews (PROSPERO) with registration number CRD420251051136.

### Study Design

The study is an overview of systematic reviews (an umbrella review) comparing the care quality (protection from infection) and the cost of using sterile gloves versus non-sterile (clean or examination) gloves in wound care and or minor surgical procedures.

### Study Selection

The study selection process for this review follows the flowchart as shown in Figure 1. Studies will be independently reviewed by two members of the research team to decide whether it should be included. The titles and abstracts will be screened first. Studies that pass this initial stage will then be assessed in full. Any disagreements will be discussed by the research team and a decision will be arrived at on the inclusion of studies. For studies that will be excluded, the reasons for exclusion will be documented.

**Fig 1.**
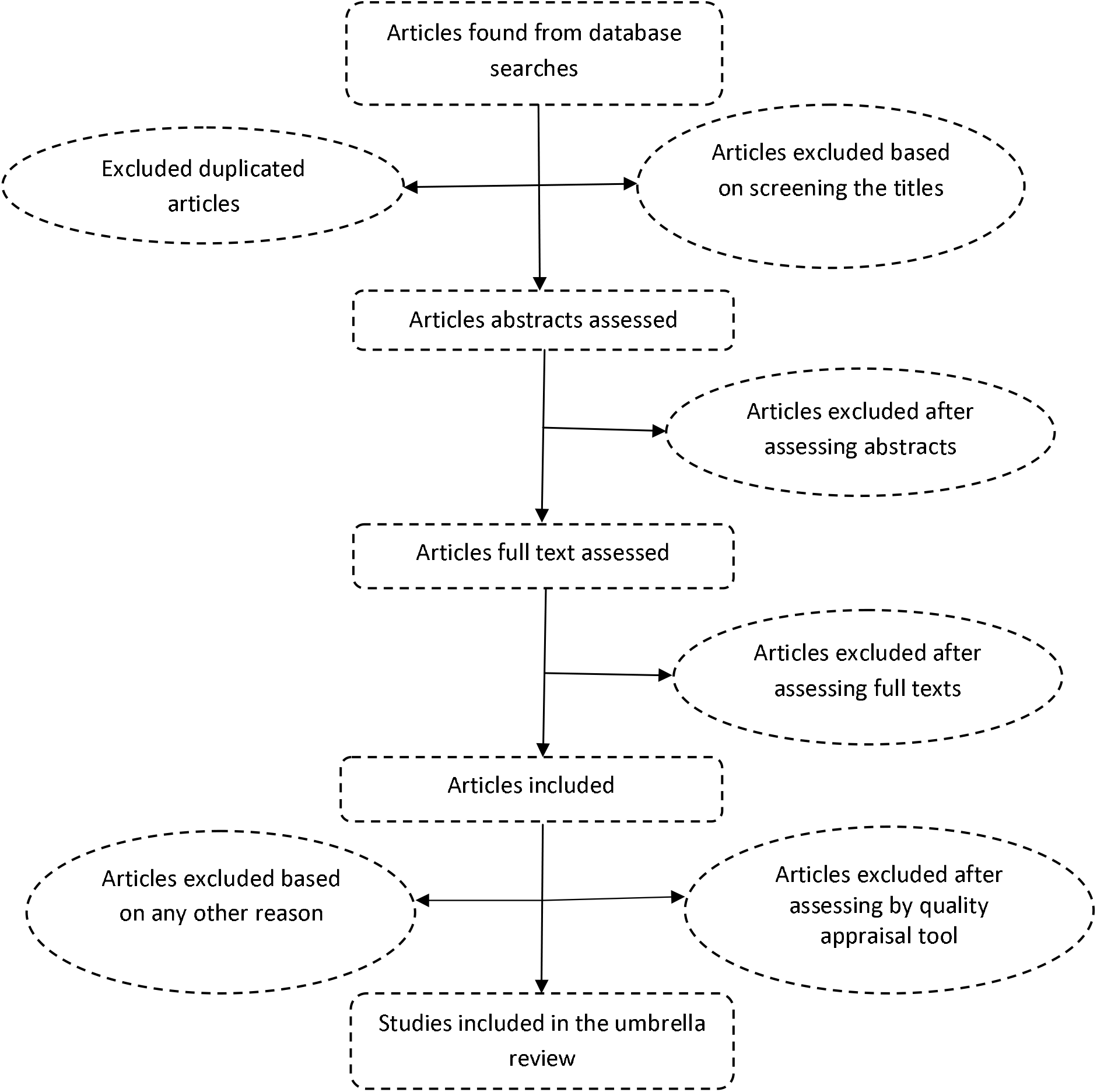
Flowchart for the selection of systematic reviews for the umbrella review.

### Eligibility Criteria

Systematic reviews will be considered for inclusion in the umbrella review if they meet the following criteria:

1. **Study type:** Systematic reviews with meta-analyses
2. **Population:** Individuals who had wound care or underwent minor surgical procedures in a healthcare setting
3. **Interventions:** Use of sterile gloves during the wound care or minor surgical procedure
4. **Comparators:** Use of examination or clean (non-sterile) gloves during wound care or minor surgical procedure
5. **Primary outcomes:** Infection rates
6. **Secondary outcomes:** Cost of glove
7. **Language:** English only

### Information Sources and Search Strategy

A comprehensive search will be conducted using PubMed, CINAHL and MEDLINE databases, covering all available systematic reviews up to May 31, 2025. Additional relevant studies found in the reference lists of the included systematic reviews will also be considered.

Search terms were developed and combined using Boolean operators ‘AND’ and ‘OR’. The keywords that will be used to search databases are; sterile gloves, surgical gloves, clean gloves, nonsterile gloves, examination gloves, wound care, wound management and wound dressing.

The final search term to be used for PubMed is:

(sterile gloves[Title/Abstract]) OR (surgical gloves[Title/Abstract]) AND (clean gloves[Title/Abstract]) OR (nonsterile gloves[Title/Abstract]) OR (examination gloves[Title/Abstract]) AND (wound care[Title/Abstract]) OR (wound dressing[Title/Abstract]) OR (wound management[Title/Abstract])

Filters applied: Systematic Review

### Data Extraction

Data will be extracted into excel spread sheet from all the included systematic reviews by two independent reviewers. The results will be compared for consistency and any concerns resolved through discussion by all authors. The information to be extracted will include the healthcare procedure reported on, department, timeline for studies included in the individual reviews, study designs reported on, outcomes measured, number of studies included in the meta-analysis for the individual systematic reviews, number of patients reported on in the meta-analysis, the outcome results, and the key findings.

### Quality Appraisal

The quality of included reviews will be assessed using the AMSTAR 2 tool. Reviews will be classified as high, moderate, low, or critically low quality. Low- and critically low-quality systematic reviews will be excluded from the study.

### Data Synthesis

A narrative synthesis will be performed. Findings will be grouped and compared based on clinical outcomes (protection from infection) and economic outcomes (cost savings). Where available, subgroup analyses by study designs or procedure involved will be highlighted.

### Ethics and Dissemination

Ethical approval is not required for this study, as it is an umbrella review. It does not involve recruiting or studying people directly. The findings will be shared at conferences and symposia, and communicated to the general public, policymakers, and stakeholders interested in the affordability of quality healthcare, and published in a peer-reviewed journal.

## Data Availability

All data produced in the present work are contained in the manuscript

## Author Contributions

Cyril E. Dzah (guarantor) conceptualized and designed the study. Joy G. L. Bulley and Valentine K. Dzah contributed to the protocol development, search strategy, and methodology. All authors reviewed and approved the final manuscript.

## References

1. Sen CK. Human wound and its burden: updated 2020 compendium of estimates. Adv wound care. 2021;10(5):281–92.

2. Farahani M, Shafiee A. Wound healing: from passive to smart dressings. Adv Healthc Mater. 2021;10(16):2100477.

3. Dinter S. THE IMPORTANCE OF EMOTIONAL INTELLIGENCE IN PATIENTS AND THEIR EFFECT ON POSTOPERATIVE WOUND HEALING. University of Rijeka. Faculty of Medicine; 2024.

4. Lei J, Sun L, Li P, Zhu C, Lin Z, Mackey V, et al. The wound dressings and their applications in wound healing and management. Heal Sci J. 2019;13(4):1–8.

5. Mihai MM, Dima MB, Dima B, Holban AM. Nanomaterials for wound healing and infection control. Materials (Basel). 2019;12(13):2176.

6. Freitas J, Lomba A, Sousa S, Gonçalves V, Brois P, Nunes E, et al. Consensus-based guidelines for best practices in the selection and use of examination gloves in healthcare settings. Nurs Reports. 2025;15(1):9.

7. Ding S, Lin F, Marshall AP, Gillespie BM. Nurses’ practice in preventing postoperative wound infections: an observational study. J Wound Care. 2017;26(1):28–37.

8. Tan YY, Chua ZX, Loo GH, Ong JSP, Lim JH, Siddiqui FJ, et al. Risk of wound infection with use of sterile versus clean gloves in wound repair at the Emergency Department: A systematic review and meta-analysis. Injury. 2023;54(11):111020.

9. Friedlander MP. The (Sterile) Gloves Are Coming Off. Clin Rev. 28:9.

10. Shrestha O, Basukala S, Bhugai N, Bohara S, Bhatt A, Thapa N, et al. Postprocedural infection rate after minor surgical procedures performed with and without sterile gloves: a systematic review and meta-analysis. Int J Surg. 2024;110(11):7341–52.

11. Wang Z, Grundy Q, Parker L, Bero L. Variations in processes for guideline adaptation: a qualitative study of World Health Organization staff experiences in implementing guidelines. BMC Public Health. 2020;20:1–13.

12. Weisz G, Nannestad B. The World Health Organization and the global standardization of medical training, a history. Global Health. 2021;17(1):96.

13. Osaki H, Skovdal M, Sørensen JB, Maaløe N, Housseine N, Dmello BS, et al. The Dilemmas and Opportunities of Co□Creating Health Interventions to Fit Local Contexts: An Ethnographic Study on the Adaptation of Clinical Guidelines in Tanzania. Heal Expect. 2024;27(5):e70073.

